# Sexually Explicit and Violent Media Use among High School Students in Vietnam: Gender-Differentiated Links with Sexual Misconduct Victimization, Perpetration, and Health

**DOI:** 10.1101/2025.04.01.25324943

**Authors:** Kathryn M. Yount, Meghan Macaulay, Kim Tu Tran

**Author notes:** Correspondence: Kathryn M. Yount, Hubert Department of Global Health, Rollins School of Public Health, 1518 Clifton Rd NE, Atlanta, GA 30322, USA.

## Abstract

**Background:** Research is lacking in lower-income countries on levels and associations of sexually explicit and sexually violent media use (SEM/SVM) with well-being outcomes in high-school students.

**Objective:** We tested theories of gendered aggression and moral incongruence to explain the associations of SEM and SVM use versus non-use with sexual misconduct perpetration/victimization and well-being outcomes among males and females attending three high schools in Ho Chi Minh City, Vietnam.

**Method:** 712 eleventh and twelfth graders were surveyed about their general health, mental health, alcohol use, beliefs about pro-violence peer norms, academic disengagement, sexual misconduct perpetration and victimization, and SEM/SVM use. Unadjusted and adjusted regression models were estimated.

**Results:** Most (79%) reported any SEM/SVM use (47% SEM only; 32% any SVM). SVM use was higher among boys (39%) than girls (27%). Among boys, users reported poorer adjusted self-rated health (SEM coef=0.43; SVM coef=0.58), higher alcohol use (SEM aOR=2.45; SVM aOR=2.35), and higher sexual misconduct perpetration (SVM aOR=5.31) than non-users. Among girls, SVM users reported higher adjusted sexual misconduct perpetration (aOR=2.81) and sexual violence victimization (aOR=5.99), and SEM users reported higher sexual misconduct victimization (aOR=2.24) than non-users.

**Conclusion:** Sexualized media use is common among high-school students in Ho Chi Minh City, especially among boys. Gendered aggression theory helps explain the effects of use on violence victimization and perpetration. Moral incongruence theory helps explain poorer health among male users. “No-phone” programs alongside staff, parent, and student outreach about the harms of sexualized media use should be tested to improve high-school students’ well-being in Vietnam.

## INTRODUCTION

*Sexual misconduct* refers to “physical or non-physical conduct of a sexual nature, including sexual harassment, stalking, dating violence, and sexual violence in the absence of clear, knowing, affirmative consent”^1^ (p. 9). Among middle- and high-school-aged students globally, experiences of sexual misconduct are common, especially for girls,^2^ and exposure is associated with multiple adverse health and social outcomes, with effects into adulthood.^3^An understudied risk factor for sexual misconduct and associated adverse conditions is exposure to sexually explicit media (SEM) or sexually violent media (SVM). *Sexually explicit media* depicts sexual intercourse, contact, or objectification, including “domination, degradation, subordination, or humiliation” most often of women^4^ and other vulnerable groups.^5^ Thus, SEM includes sexualized media that depicts individuals attempting to inflict or inflicting psychological (but not physical) harm on others. *Sexually violent media* is SEM that depicts individuals attempting to inflict or inflicting physical harm on others.^5^ Meta-analyses have found higher rape-myth acceptance with SEM use and SVM use, and higher aggression with SEM use ((r =.15 [.13, .17]) and especially SVM use (r =.25 [.19, .31]).^5^ Yet, both meta-analyses included mostly male samples from higher-income Western settings,^5,6^ limiting tests of gender-differentiated effects and inferences to lower-income, non-Western settings.

Research on the effects of SEM and SVM use in young people is rare in low- and middle-income countries (LMICs).^5-7^ In Vietnam, undergraduate men have reported high rates of any SEM use (86%), any SVM use (41%), and at least monthly SVM use (17%) in the prior six months.^7^ Men reporting any or at least monthly SVM use had 2-4 times higher adjusted odds than their counterparts of sexually violent behavior.^7^ Similar research among high-school students is even rarer in LMICs. Among eleventh and twelfth graders in three high schools in Ho Chi Minh City, Vietnam, a majority (55%) have experienced some form of sexual misconduct since attending high school;^8^ yet, the relationship of SEM and SVM use with sexual misconduct and other well-being outcomes was not assessed.

Here, we analyzed data from the above-mentioned study of students in Ho Chi Minh City to test theories of gendered aggression and moral incongruence to explain associations of SEM and SVM use with sexual misconduct victimization and perpetration; self-reported health and risk behaviors; academic disengagement; and beliefs in pro-violence peer norms. Findings offer insights about interventions in high schools to address gender-differentiated harms of SEM and SVM use in Vietnam, and perhaps beyond.

## BACKGROUND

### Sexual Misconduct and Well-Being among Young People

A global systematic review of sexual misconduct in middle- and high-school-aged students found that prevalence rates of exposure to specific forms of sexual harassment were higher among female than male students (9% to 47% versus 12% to 35%), as were prevalence rates of sexual assault (7% to 12% versus 2% to 6%), any dating violence (21% versus 10%), physical dating violence (11% to 30% versus 14%), and sexual dating violence (8% versus 3%).^2^ Exposure to sexual misconduct at this age has been associated with poorer mental health, self-harm and suicide ideation and attempts, drug and alcohol use, aggression and violence, poor sexual health, and poor academic performance.^2,3^

### Effects of Sexualized Media Use on Sexual Misconduct: *Gendered* Aggression

An understudied risk factor for incidents of sexual misconduct is the use of SEM and/or SVM. Mimicry, sexual scripts, social cognition, and cultivation theories are relevant frameworks.^9,10^ Mimicry refers to imitating behaviors, including those seen in media.^10^ Sexual script theory asserts that individuals model their sexual expectations, norms, desires, and actions after portrayals of sex in their culture, such as in media.^11^ Social cognitive theory employs the concept of observational learning,^12^ including about sexual activity and violence from media.^10^ Young people routinely employ observational learning,^13^ including in cases of observed violence.^14^ Observational learning often occurs over time, with enactment continuing beyond the observed event,^13^ Cultivation theory, also applied to media-violence research,^6^ posits that greater and repeated use of media contributes to greater acceptance of the norms and beliefs it conveys.^15^ The repeated observation of arousing and violent content can cause desensitization, diminishing negative affect when viewing or enacting violence.^10^

The *general aggression* meta-theory incorporates these theories of aggression,^16^ and a modified interpretation of this model links SEM and SVM use with higher risks of sexual misconduct perpetration by men *and* sexual misconduct victimization of women. According to the general model, behavior is based on the learning, activation, and application of related knowledge structures, such as scripts stored in memory that may be learned from observing characters in media.^17^ Once learned, the script helps individuals understand similar situations and guides behavior. One first selects a script from memory to represent the situation, assumes a role in the script, and behaves accordingly.^18^ As these knowledge structures are practiced, they may become more complex, differentiated, and resistant to change.

Media contain many reinforcing scripts of how men should treat women. In “mainstream pornography” in the U.S., sex scenes often portray verbal (49%) and physical (88%) aggression, usually by men against women.^19,20^ As such, SEM and SVM may lead the user to objectify women and accept violence against them.^21^ When sexual scripts in media largely depict men’s objectification of and verbal aggression toward women (in SEM), men’s physical aggression toward women (in SVM), and women’s subordination to men (in SEM and SVM), a *gendered aggression model* would predict that: men who repeatedly use such content will become more habitually violent, and women who repeatedly use such content will become more habitually victimized because both users tend to assume the role of their gender in the script.

Evidence for this hypothesis remains inconclusive. In a convenience sample of mainly white college students in the U.S., SEM-use frequency was positively associated with aggressor, target, and uncommon/degrading sexual behavior for men and women.^22^ In global meta-analyses, SEM use and SVM use have shown positive effects on rape-myth acceptance^5,6^ and aggression (SEM use r =.15 (.13, .17); SVM use r =.25 (.19, .31)) and similar effect sizes across gender, age, and student/non-student samples.^5^ Yet, these analyses could not test gender-differentiated effects of SEM and SVM use on aggression due to insufficient female samples or generalize findings “globally” because most studies came from higher-income, Western settings.^5^

### Effects of Sexualized Media Use on Health: *Moral Incongruence*

Other literature has examined the effects of SEM use and SVM use on broader measures of adolescent psychosocial well-being.^23^ One consistent finding is that adolescent SEM users more often exhibit emotional problems.^24^ Some researchers argue that individuals suffering from these problems use SEM as a coping strategy; whereas, others propose that SEM use may contribute to psychological distress.^25^ Extending the second argument, the link between SEM use and emotional problems may *depend on* the congruence between a user’s moral beliefs about SEM and their use practice.^25^ SEM/SVM use has risen more quickly than its approval, and more young users may experience an incongruence between their moral beliefs and behaviors related to SEM/SVM use. Thus, the association between SEM and SVM use and psychosocial well-being may depend on the extent of the user’s cognitive dissonance—the psychological distress felt when stated values or beliefs do not align with actions. Social values about what is “good” or “bad” may influence the stress-formation process because these values define how actors interpret their behavior.^26^ This research comes mostly from higher-income Western countries, and to our knowledge, the model has not been tested among adolescents in LMICs.

### Gendered Aggression and Moral Incongruence in Vietnam

A gendered aggression model for the effects of SEM and SVM use may be salient in Vietnam, where historical and prevailing gender norms reinforce gendered sexual scripts in media. In Vietnam’s vertical patrilineal kinship system, descent is traced through fathers,^27^ men are viewed as superior to women,^27^ and senior men have been designated as “pillars” of the household.^27^ Confucianism aligns femininity with compliance and endurance to uphold family harmony and masculinity with “hot anger” and violence.^27^

Gendered scripts in mainstream and sexualized media reinforce these gender norms. For example, 25% of 400 news-media samples have mentioned domestic violence, more often depicting women as victims and men as perpetrators, and more often depicting women in stereotypically domestic roles.^28^ Seventy studies in Asia, including three in Vietnam, have addressed the phenomenon of *deep-fake porn*, in which the creators use artificial intelligence to overlay mostly women’s faces onto sexually explicit or violent images or videos.^29^ Increases in *problematic internet use* among 18-25 year-olds in recent years may have increased exposure to these gendered sexual scripts.^30^ Thus, observing gendered scripts in daily life and gendered sexual scripts in various media may increase a male viewer’s tendency to enact sexual misconduct and a female viewer’s tendency to experience it.^31^

A moral incongruence model also may be salient in Vietnam, where the State has defined commercial sex and parents have defined teenage sexuality as “social evils”,^32^ while premarital sex among young people has been increasing.^33^ Consequently, young people are interpreting their behavior amidst parental expectations of premarital chastity, especially for girls,^32,34^ peer expectations for sex, and prevailing norms of feminine obedience, masculine dominance, and men’s entitlement to sex.^31,34^ A young person’s interpretation of their behavior against this complex social value system may contribute to “stress valuation” among SEM/SVM users, leading to poorer well-being, especially among boys, who are more frequent users of sexualized media.^24^

First-year undergraduate men in Vietnam report high rates of any SEM use (86%), any SVM use (41%), and SVM use at least monthly (17%) in the prior six months.^7^ Men exposed to any or at least monthly SVM have had 2-4 times higher adjusted odds than their counterparts of engaging in sexually violent behavior.^7^ Among eleventh and twelfth graders attending three high schools in Ho Chi Minh City, 55% reported experiencing any sexual misconduct since starting high school.^8^ The prevalence of sexual harassment victimization during that period was 40% perpetrated by staff and 30% perpetrated by students. Also in that period, the prevalence of stalking victimization was 18%, and the prevalence of physical or emotional dating violence victimization was 13%.^8^ Nearly one in ten students reported any sexual violence victimization. Boys reported more often than girls unwanted sexual attention (19% versus 11%) and sexual coercion (10% versus 5%) *by staff*. Girls reported more often than boys physical and emotional dating violence victimization (15% versus 10%). The authors did not test gender-differentiated effects of SEM and SVM use on sexual misconduct victimization, perpetration, or broader psychosocial well-being.

### Hypotheses

Using novel data from high school students in Ho Chi Minh City, we tested the associations of SEM use and SVM use with sexual misconduct victimization and perpetration, self-rated general health and mental health, alcohol use, academic disengagement, and perceived pro-violence norms among peers. Based on the general and gendered aggression model, we expected that SEM use and SVM use will be associated with:

(1) higher sexual misconduct victimization and perpetration in the pooled sample due to
(2) higher sexual misconduct victimization in girls and higher sexual misconduct perpetration in boys.

Based on the moral incongruence model, we expected that:

(3) SEM use and SVM use will be associated with poorer self-rated health, poorer mental health, lower quality of life, and alcohol use;
(4) beliefs that violence against women is accepted among peers will strengthen this association; and
(5) these direct and moderating associations will be stronger among boys than girls.

## METHOD

### Ethical Considerations

The study received ethical approval from the Institutional Review Board of the University of Medicine and Pharmacy at Ho Chi Minh City (3746/DHYD-HDDD).

### Sample

Three high schools in Ho Chi Minh City were selected randomly from high-, middle-, and low-ranked school strata, defined by students’ scores on the national entrance test. This selection ensured a demographically diverse sample of students. Eleventh- and twelfth-graders attending selected high schools were eligible, which ensured that exposures and outcomes covered the periods of high-school attendance only.

### Data Collection

The questionnaire included modules from the GlobalConsent efficacy trial^35^ and the Administrative Researcher Campus Climate Collaborative (ARC3) survey.^1^ The GlobalConsent online survey included nine modules with scales validated in Western populations that were translated, cognitively tested, adapted, and validated among undergraduate men in Vietnam.^35-37^ The module on media use,^7,37^ adapted from the Kids Online survey,^38^ provided the exposure measures for the present study.

The ARC3 survey, a free campus-climate survey designed by sexual assault researchers and administrators for U.S. institutions of higher education, assesses forms of sexual misconduct identified as Title IX violations. The survey includes 19 modules covering types of sexual misconduct since enrollment, climate-related factors, and possible outcomes of sexual misconduct.^1^ Included measures have shown strong psychometric properties in prior research, ARC3 pilot research, and subsequent research at seven U.S. universities.^1,39^ The ARC3 survey provided the outcome measures for the present study. Local experts reviewed the modules for feasibility and suitability with high-school students in Vietnam.

The final questionnaire included 20 modules. A demographics module included questions on sex at birth, gender identity, and sexual orientation, ethnicity, age, grade, organizational membership, and living arrangements.^1,37^ The SEM/SVM module asked about the frequency of using five types of sexually explicit material (text-based, partial nudity, full nudity, non-violent sexual acts, violent sexual acts) through four types of media (internet, TV/movie/DVD, book/magazine, social media).^37^ The outcomes modules included measures of sexual harassment victimization by faculty/staff and students,^40^ stalking victimization and perpetration,^41^ dating violence victimization and perpetration,^42,43^ sexual violence victimization and perpetration,^44^ perceived peer norms about sex, dating violence, and sexual violence,^45^ self-rated health and general mental health,^46^ satisfaction with life,^47^ and academic disengagement.^48,49^ More sensitive questions on sexual misconduct were asked in later modules. The team developed a customized and secure online survey using REDCap.^50^

Parents were invited to provide written informed consent for their children to participate. Students were invited to provide written informed assent to participate. Assenting students whose parents consented convened in classrooms and received a link to the online survey through their mobile phones with 3G connectivity. The survey was anonymous, meaning no identifiers were collected, and a randomly generated identification number was assigned to each participant. Two non-study staff were present in the classroom to assist participants with any technological issues that students encountered while entering and completing the survey. The survey-link address was altered after each data collection to prevent students from re-accessing the system. Students who completed the survey received a $3 voucher.

### Outcomes

*Self-rated health* was measured with one question about perceived health (0=Excellent to 4=Poor). *Mental health* was measured as the mean frequency (0=Never to 4=Always) in the prior four weeks of experiencing five mental states, such as feeling very nervous. Items were coded so higher scores denoted worse mental health (α=0.71 in this sample). *Self-reported satisfaction with life* was measured with one question about perceived satisfaction with life (0=Strongly Agree to 4=Strongly Disagree). *Alcohol consumption* was measured as the frequency (0=Never to 4=four or more times a week) at the time of the survey of consuming alcohol, and response options were recoded to capture any (=1) versus no (=0) alcohol consumption. *Academic disengagement* was measured as the mean frequency (0=Almost Never to 4=Almost Always) in the prior semester of eight behaviors, such as missing class. Higher scores denoted higher academic disengagement (α=0.81 in this sample).

*Sexual misconduct perpetration* was measured as reporting to have ever (versus never) committed, since attending high school, any of six physical dating violence items (e.g., “I hit the person”), any of 10 stalking items (e.g., “made unwanted calls to them”), or any of 25 act-by-tactic sexual violence items (e.g., “had oral sex with someone…without their consent”). Internal consistency of the items was 0.97 in this sample. *Sexual violence perpetration* was measured as reporting to have committed any of the 25 act-by-tactic items on sexual violence since attending high school (α=0.98 in this sample).

*Sexual misconduct victimization* was measured as having ever (versus never) reported experiencing, since attending high school, any of 16 sexual harassment items by faculty/staff or students (e.g., “treated you differently because of your sex”), any of six physical dating violence items (e.g., “hit you”), any of 10 stalking items (e.g., “made unwanted phone calls to you”), or any of 25 act-by-tactic sexual violence items (e.g., “had oral sex with you…without your consent”). Internal consistency of the items was 0.98 in this sample. *Sexual violence victimization* was measured as having ever (versus never) reported experiencing any of the 25 act-by-tactic items on sexual violence since attending high school (α=0.96 in this sample).

### Exposure

Use of sexualized media was measured categorically as no use (=0), any SEM use only (=1), and any SVM use (=2) in the prior six months. SVM was defined as “images or video of a woman performing a sex act in which she was choked, hit, humiliated, or forced.”

### Moderator

The moderator variable was *beliefs about pro-violence peer norms*. It was measured as the mean response to seven questions about the participant’s level of agreement (0=Strongly Disagree to 4=Strongly Agree) that his or her friends would approve of several behaviors, such as having many sexual partners, insulting or swearing at dates, and forcing someone to have sex. Higher scores denoted higher perceived peer support for risky and violent sexual behaviors (α=0.89 in the sample).

### Covariates

Covariates measured school ranking (low, middle, high), grade level (eleventh [reference], twelfth), gender identity (female [reference], male), sexual orientation (heterosexual [reference], non-heterosexual), living situation (without parents [reference], with parents), feeling safe on campus (Agree [reference]; Neutral; Disagree), and perceived school climate (Likely [reference]; Neutral; Unlikely to support victims).

### Analysis

First, we performed descriptive analysis to understand the unweighted distributions of outcomes, exposures, and demographic covariates and to identify potential confounders of the relationships of interest. We examined the distributions of covariates and the SEM/SVM exposure for the pooled sample and tested for differences in covariates across media-use groups (none, SEM only, any SVM). For categorical covariates, we computed and reported the exact p-value for the χ^2^ test statistic for differences in their distributions across media-use groups. For outcomes, we estimated mean scores for continuous measures and prevalence rates for binary measures in the full sample. We performed bivariate analysis of variance (ANOVA) to test for differences in scores for continuous outcomes across media-use groups, and we computed the χ2 statistic to test for differences in the prevalence rates of binary outcomes across media-use groups.

Second, we estimated linear regressions for continuous outcomes and logistic regressions for binary outcomes to test for their associations with SEM use and SVM use, relative to no use. For each outcome, we estimated a bivariate model and a multivariate model adjusted for all covariates using the full sample, female-only sample, and male-only sample. Unadjusted and adjusted coefficients for linear regressions and odds ratios for logistic regressions were presented with their 95% confidence intervals. To test the general aggression model, lower bounds >1.00 for adjusted odds ratios for the associations of SEM use and SVM use with sexual misconduct and sexual violence perpetration and victimization in the pooled sample provided tests for hypothesis 1. To test a gendered aggression model, lower bounds >1.00 for adjusted odds ratios for the associations of SEM use and SVM use with sexual misconduct and sexual violence victimization in the female sample and with sexual misconduct and sexual violence perpetration in the male sample provided tests for hypothesis 2. To test incongruence theory, lower bounds >0.00 for adjusted coefficients and >1.00 for adjusted odds ratios for the associations of SEM use and SVM use with health outcomes provided tests for hypothesis 3.

Finally, we added a measure for participants’ beliefs about pro-violence peer norms, a beliefs-by-SEM-use interaction term, and a beliefs-by-SVM-use interaction term to all adjusted regression models for health outcomes. Lower bounds of the confidence interval for these interaction terms >0.00 in the pooled sample and the male-only sample provide tests for hypotheses 4-5.

## RESULTS

### Sample Characteristics

Approximately 20.5% of participants had not used any SEM in the prior six months (Table 1). Just under half (47.3%) had used SEM only in that period, and almost one-third (32.2%) had used SVM. Among female students, 24.0% had never used any SEM in the prior six months, 48.7% had used SEM only, and 27.3% had used SVM. Among male students, 15.4% had never used any SEM in the prior six months, 45.5% had used SEM, and 39.2% had used SVM.

**Table 1.**
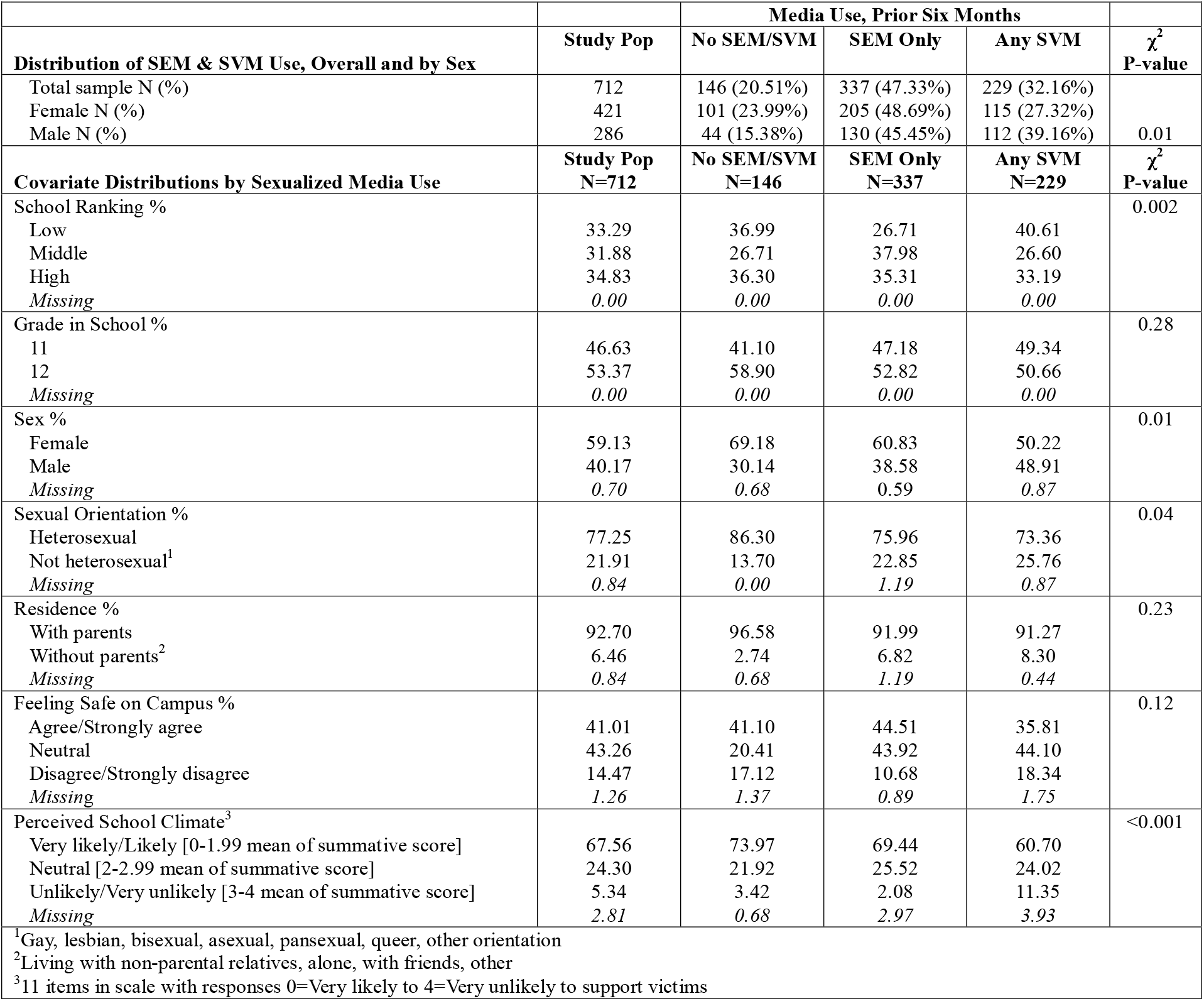
Sample Characteristics (Unweighted): Eleventh and Twelfth Graders Attending Three High Schools in Ho Chi Minh City, Vietnam, 2023.

The pooled sample was distributed evenly across the three schools (Table 1); however, a higher percentage of participants attending the lower-ranked school was represented among SVM users (40.6%) than among SEM users (26.7%) or non-users (37.0%). The sample was approximately evenly distributed across grades (46.6% eleventh graders; 53.4% twelfth graders), and this distribution did not differ across media-user groups. Overall, a higher percentage of participants identified as female (59.1%) than as male (40.2%), and a higher percentage of females were represented among those who had not used SEM/SVM in the prior six months (69.2%) than those who had used SEM (60.8%) or SVM (50.2%). About 21.9% of the sample self-identified as non-heterosexual (gay, lesbian, bisexual, asexual, pansexual, queer, or other orientation), and these participants were represented more often among SVM users (25.8%) than among non-users (13.7%) in the prior six months. Living without parents was uncommon overall (6.5%), and this percentage did not differ across media-user groups. A minority (41.0%) of participants agreed that they felt safe on campus, and this percentage did not differ across media-user groups. A majority of participants felt it likely that their school would support victims of sexual misconduct, and a higher percentage of SVM users (11.4%) than SEM users (2.1%) and non-users (3.4%) thought school support for victims was unlikely.

### Social and Health Status by Media Use

In the pooled sample, mean self-rated health was 1.65 or “above average,” and did not differ by user group (Table 2). On average, participants “agreed” or were “neutral” about their life satisfaction (mean 1.71); however, life satisfaction was more “neutral” among SVM users (mean 1.85). On average, participants “rarely” or “sometimes” experienced symptoms of anxiety or depression in the prior four weeks (mean 1.75), and SVM users reported symptoms more often (mean 1.84). Over one-third (34.6%) of participants reported consuming any alcohol, and a dose-response relationship was observed between media use and alcohol consumption (non-users: 27.6%; SEM users: 33.3%; SVM users: 41.2%).

**Table 2.** Outcomes, Overall and by Media Use, Students Attending Three High Schools in Ho Chi Minh City, Vietnam, 2023.

|  | **Study Population** | **Media Use, Prior Six Months** |  |  | **ANOVA or X<sup>2</sup> P-value** |
| --- | --- | --- | --- | --- | --- |
|  |  | **No SEM/SVM** | **SEM Use Only** | **Any SVM Use** |  |
| **Outcomes (Unweighted)** | Mean (SD) or N (%) | Mean (SD) or N (%) | Mean (SD) or N (%) | Mean (SD) or N (%) |  |
| Self-rated Health [0=excellent to 4=poor] *N*=708 | 1.65 (1.05) | 1.51 (0.99) | 1.68 (1.03) | 1.70 (1.12) | **0.17** |
| Satisfied with Life [0=str agree to 4=str disagree] *N*=706 | 1.71 (0.98) | 1.60 (0.98) | 1.66 (0.94) | 1.85 (1.04) | **0.03** |
| Depressive and Anxiety Symptoms<sup>1</sup> *N*=699 | 1.75 (0.68) | 1.71 (0.74) | 1.71 (0.66) | 1.84 (0.67) | **0.05** |
| Any Alcohol Use *N*=710 | 246 (34.55%) | 40 (27.59%) | 112 (33.33%) | 94 (41.23%) | 0.02 |
| Academic Disengagement<sup>2</sup> *N*=697 | 0.85 (0.64) | 0.78 (0.70) | 0.82 (0.54) | 0.94 (0.72) | **0.03** |
| Beliefs in Pro-Violence Peer Norms<sup>3</sup> *N*=696 | 0.57 (0.59) | 0.51 (0.60) | 0.52 (0.54) | 0.68 (0.65) | **0.01** |
| Any sexual-misconduct perpetration<sup>4</sup> *N*=712 | 128 (17.98%) | 13 (8.90%) | 55 (16.32%) | 60 (26.20%) | <0.001 |
| Any sexual-violence perpetration<sup>4</sup> *N*=709 | 34 (4.80%) | 1 (0.68%) | 9 (2.69%) | 24 (10.53%) | <0.001 |
| Any sexual-misconduct victimization<sup>4</sup> *N*=651 | 432 (66.36%) | 70 (52.24%) | 214 (70.86%) | 148 (68.84%) | <0.001 |
| Any sexual-violence victimization<sup>4</sup> *N*=646 | 65 (10.06%) | 4 (2.84%) | 24 (8.03%) | 37 (17.96%) | <0.001 |

Notes. Mean (SD) for outcomes on 0-4 scale, n (%) for binary outcomes. P-value based on X2 statistic for binary outcomes, ANOVA for means.
<sup>1</sup>Mean of 5 items each scored 0=never, 4=always
<sup>2</sup>Mean of 8 items each scored 0=almost never, 4=almost always
<sup>3</sup>Mean of 12 items each scored 0=strongly disagree, 4=strongly agree
<sup>4</sup>Since enrollment

The mean score for academic disengagement was 0.85 overall, suggesting that these practices were uncommon among participants. However, the mean score for academic disengagement was higher among SVM users (0.94) than among SEM users (0.82) or non-users (0.78). Similarly, participants tended to disagree with statements reflecting pro-violence peer norms (mean score 0.57); yet, the mean score for beliefs in pro-violence peer norms was higher among SVM users (0.68) than among SEM users (0.52) and non-users (0.51).

The prevalence of any sexual misconduct perpetration was 18.0% overall and was highest among SVM users (26.2%), followed by SEM users (16.3%) and non-users (8.9%). The prevalence rate of sexual violence perpetration was 4.8% overall and was higher among SVM users (10.5%) than among SEM users (2.7%) and non-users (0.7%). The prevalence of sexual misconduct victimization was 66.4% overall and was higher among SEM users (70.1%) and SVM users (68.8%) than among non-users (52.2%). The prevalence rate of sexual violence victimization was 10.1% overall and was higher among SVM users (18.0%) than among SEM users (8.0%) and non-users (2.8%).

### Regression Results

In the pooled sample, neither SEM nor SVM use in the prior six months was associated with self-rated health or general mental health (Table 3). The unadjusted association of SVM use with poorer satisfaction with life (coef.=0.24 (0.04, 0.45)) was fully attenuated in adjusted models. Compared to non-use, prior SVM use was associated with 1.8 times higher odds of any alcohol use (uOR=1.84 (1.17, 2.89); aOR=1.80 (1.12, 2.90)) and positively associated with academic disengagement (coef.=0.16 (0.03, 0.29); coef.=0.14 (0.004, 0.27)). The unadjusted association of prior SVM use with beliefs in pro-violence peer norms (coef.=0.16 (0.03, 0.29)) was fully attenuated in adjusted models. Finally, compared to non-use in adjusted models, prior SVM use was associated with 3.3 times higher odds of sexual misconduct perpetration, 11.8 times higher odds of sexual violence perpetration, 2.2 times higher odds of sexual misconduct victimization, and 7.6 times higher odds of sexual violence victimization (Table 3). Compared to non-use in adjusted models, SEM use was associated with 2.1 times higher odds of sexual misconduct victimization.

**Table 3.** Associations of Health, Academic, Normative, and Behavioral Outcomes with Media Use in the Prior Six Months, Students Attending Three High Schools in Ho Chi Minh City, Vietnam, 2023.

| Outcome<br>Exposure: SEM/SVM use prior 6 mo (ref: none) | Unadjusted Odds Ratio/Regression Coefficient |  |  |  |  |  | Adjusted<sup>6</sup> Odds Ratio/Regression Coefficient |  |  |  |  |  |
| --- | --- | --- | --- | --- | --- | --- | --- | --- | --- | --- | --- | --- |
|  | Total Sample |  | Females |  | Males |  | Total Sample |  | Females |  | Males |  |
|  | OR/Coef | 95% CI | OR/Coef | 95% CI | OR/Coef | 95% CI | OR/Coef | 95% CI | OR/Coef | 95% CI | OR/Coef | 95% CI |
| **Self-Rated Health (0=excellent to 4=poor)<sup>1</sup>** |  |  |  |  |  |  |  |  |  |  |  |  |
| SEM use | 0.17 | -0.03, 0.38 | 0.09 | -0.16, 0.34 | **0.43** | **0.08, 0.79** | 0.20 | -0.01, 0.40 | 0.07 | -0.18, 0.33 | **0.43** | **0.09, 0.77** |
| SVM use | 0.20 | -0.02, 0.42 | -0.01 | -0.29, 0.27 | **0.64** | **0.28, 1.00** | 0.21 | -0.01, 0.43 | -0.04 | -0.33, 0.26 | **0.58** | **0.23, 0.93** |
| **Satisfied with Life (0=strongly agree to 4=strongly disagree)<sup>1</sup>** |  |  |  |  |  |  |  |  |  |  |  |  |
| SEM use | 0.06 | -0.13, 0.25 | -0.02 | -0.25, 0.21 | 0.21 | -0.14, 0.55 | 0.05 | -0.14, 0.24 | -0.03 | -0.26, 0.19 | 0.24 | -0.10, 0.58 |
| SVM use | **0.24** | **0.04, 0.45** | 0.19 | -0.06, 0.45 | 0.30 | -0.05, 0.65 | 0.14 | -0.07, 0.34 | 0.11 | -0.16, 0.37 | 0.22 | -0.13, 0.57 |
| **Depressive and Anxiety Symptoms<sup>1,2</sup>** |  |  |  |  |  |  |  |  |  |  |  |  |
| SEM use | -0.001 | -0.13, 0.13 | -0.04 | -0.21, 0.13 | 0.11 | -0.11, 0.33 | -0.001 | -0.13, 0.13 | -0.03 | -0.20, 0.14 | 0.07 | -0.15, 0.29 |
| SVM use | 0.13 | -0.01, 0.28 | 0.12 | -0.07, 0.31 | 0.22 | -0.01, 0.44 | 0.09 | -0.06, 0.23 | 0.07 | -0.12, 0.27 | 0.14 | -0.09, 0.36 |
| **Any alcohol use at time of survey (ref: never)<sup>1</sup>** |  |  |  |  |  |  |  |  |  |  |  |  |
| SEM use | **1.31** | **0.85, 2.01** | 0.97 | 0.57, 1.63 | 2.13 | 0.97, 4.68 | 1.27 | 0.81, 1.98 | 0.89 | 0.52, 1.55 | **2.45** | **1.07, 5.62** |
| SVM use | **1.84** | **1.17, 2.89** | 1.76 | 0.999, 3.10 | 2.20 | 0.99, 4.90 | **1.80** | **1.12, 2.90** | 1.69 | 0.93, 3.08 | **2.35** | **1.01, 5.46** |
| **Academic Disengagement<sup>1,3</sup>** |  |  |  |  |  |  |  |  |  |  |  |  |
| SEM use | 0.04 | -0.09, 0.16 | 0.04 | -0.11, 0.18 | 0.04 | -0.21, 0.29 | 0.02 | -0.10, 0.14 | 0.04 | -0.10, 0.17 | -0.03 | -0.27, 0.20 |
| SVM use | **0.16** | **0.03, 0.29** | **0.16** | **0.004, 0.32** | 0.18 | -0.08, 0.43 | **0.14** | **0.004, 0.27** | 0.14 | -0.02, 0.30 | 0.09 | -0.16, 0.33 |
| **Beliefs in Pro-violence Peer norms<sup>1,4</sup>** |  |  |  |  |  |  |  |  |  |  |  |  |
| SEM use | 0.01 | -0.11, 0.13 | 0.03 | -0.09, 0.15 | -0.06 | -0.30, 0.17 | -0.02 | -0.14, 0.10 | 0.01 | -0.12, 0.13 | -0.08 | -0.32, 0.16 |
| SVM use | **0.16** | **0.04, 0.29** | 0.13 | -0.01, 0.27 | 0.12 | -0.12, 0.36 | 0.11 | -0.02, 0.23 | 0.10 | -0.04, 0.25 | 0.09 | -0.15, 0.33 |
| **Any sexual misconduct perpetration<sup>1,5</sup> (ref: no)** |  |  |  |  |  |  |  |  |  |  |  |  |
| SEM use | **2.00** | **1.05, 3.78** | 1.44 | 0.67, 3.09 | 3.42 | 0.98, 11.91 | 1.60 | 0.82, 3.12 | 1.21 | 0.54, 2.69 | 2.68 | 0.73, 9.86 |
| SVM use | **3.63** | **1.91, 6.90** | 2.79 | 1.28, 6.11 | 5.71 | 1.65, 19.74 | **3.29** | **1.68, 6.45** | **2.81** | **1.23, 6.40** | **5.31** | **1.45, 19.41** |
| **Any sexual violence perpetration<sup>1,5</sup> (ref: no)** |  |  |  |  |  |  |  |  |  |  |  |  |
| SEM use | **4.00** | **0.50, 31.89** | 0.99 | 0.09, 11.00 | 0.23 | 0.09, 0.62 | 2.76 | 0.33, 22.98 | 0.99 | 0.89, 11.46 | **0.22** | **0.08, 0.61** |
| SVM use | **17.06** | **2.28, 127.53** | 4.55 | 0.52, 39.57 | 1.00 | omitted | **11.78** | **1.53, 91.04** | 3.51 | 0.36, 34.67 | 1.00 | omitted |
| **Any sexual misconduct victimization<sup>1,5</sup> (ref: no)** |  |  |  |  |  |  |  |  |  |  |  |  |
| SEM use | **2.22** | **1.46, 3.38** | 2.48 | 1.47, 4.17 | 2.02 | 0.97, 4.23 | **2.11** | **1.34, 3.32** | **2.24** | **1.28, 3.92** | 1.87 | 0.83, 4.23 |
| SVM use | **2.02** | **1.29, 3.15** | 1.71 | 0.98, 3.01 | 2.74 | 1.28, 5.89 | **2.15** | **1.32, 3.52** | 1.75 | 0.94, 3.28 | **2.80** | **1.20, 6.56** |
| **Any sexual violence victimization<sup>1,5</sup> (ref: no)** |  |  |  |  |  |  |  |  |  |  |  |  |
| SEM use | **2.99** | **1.02, 8.79** | 1.69 | 0.54, 5.31 | 0.59 | 0.25, 1.38 | 2.81 | 0.94, 8.46 | 1.52 | 0.47, 4.94 | 0.58 | 0.23, 1.45 |
| SVM use | **7.50** | **2.61, 21.56** | 6.00 | 1.99, 18.10 | 1.00 | omitted | **7.64** | **2.57, 22.75** | **5.99** | **1.88, 19.06** | 1.00 | omitted |

<sup>1</sup>Linear regressions used for outcomes on mean or 0-4 scale, logistic regression for binary outcomes. <sup>2</sup>Mean response to 5 items 0=never to 4=always <sup>3</sup>Mean response to 8 items 0=almost never to 4=almost always <sup>4</sup>Mean response to 12 items 0=strongly disagree to 4=strongly agree <sup>5</sup>Since enrolling in high school <sup>6</sup>School ranking (low, middle, high), grade (11<sup>th</sup> [reference], 12<sup>th</sup>), sex (female [reference], male), sexual orientation (heterosexual [reference], non-heterosexual), residence (without [reference], with parents), feeling safe on campus (Agree; Neutral; Disagree), and perceived school climate (Likely; Neutral; Unlikely to support victims)

Several gender-differentiated associations were observed between media use and outcomes (Table 3). Among boys, in adjusted models, SEM use and SVM use, respectively, were associated with poorer self-rated health (coef.=0.43 (0.09, 0.77); coef.=0.58 (0.23, 0.93)) and over two times higher odds of any alcohol use (aOR=2.45 (1.07, 5.62); aOR=2.35 (1.01, 5.46)). Any SVM use also was associated with more than five times higher adjusted odds of any sexual misconduct perpetration (aOR=5.31 (1.45, 19.41)) and almost three times higher adjusted odds of sexual misconduct victimization (aOR=2.80 (1.20, 6.56)).

Among girls, SVM use was associated with almost three times higher adjusted odds of sexual misconduct perpetration (aOR=2.81 (1.23, 6.40)) and six times higher adjusted odds of sexual violence victimization (aOR=5.99 (1.88, 19.06)). SEM use was associated with over two times higher adjusted odds of sexual misconduct victimization (aOR=2.24 (1.28, 3.92)). Neither form of media use was associated with other health or social outcomes among female students (Table 3).

Finally, several interactions between SEM use and SVM use with beliefs in pro-violence peer norms were significant, especially among boys (Table 4). In that group, stronger beliefs that peers accepted violence against women strengthened the association of SEM use with poorer self-rated health and poorer mental health and SVM use with poorer mental health and increased academic disengagement. Among girls, no significant interactions were observed between beliefs about pro-violence peer norms and SVM use, and having stronger beliefs that peers accepted violence against women did not alter the association of SEM use with three well-being outcomes. For the remaining two well-being outcomes (poor mental health and academic disengagement), significant interactions of beliefs and SEM use had the opposite valence.

**Table 4.**
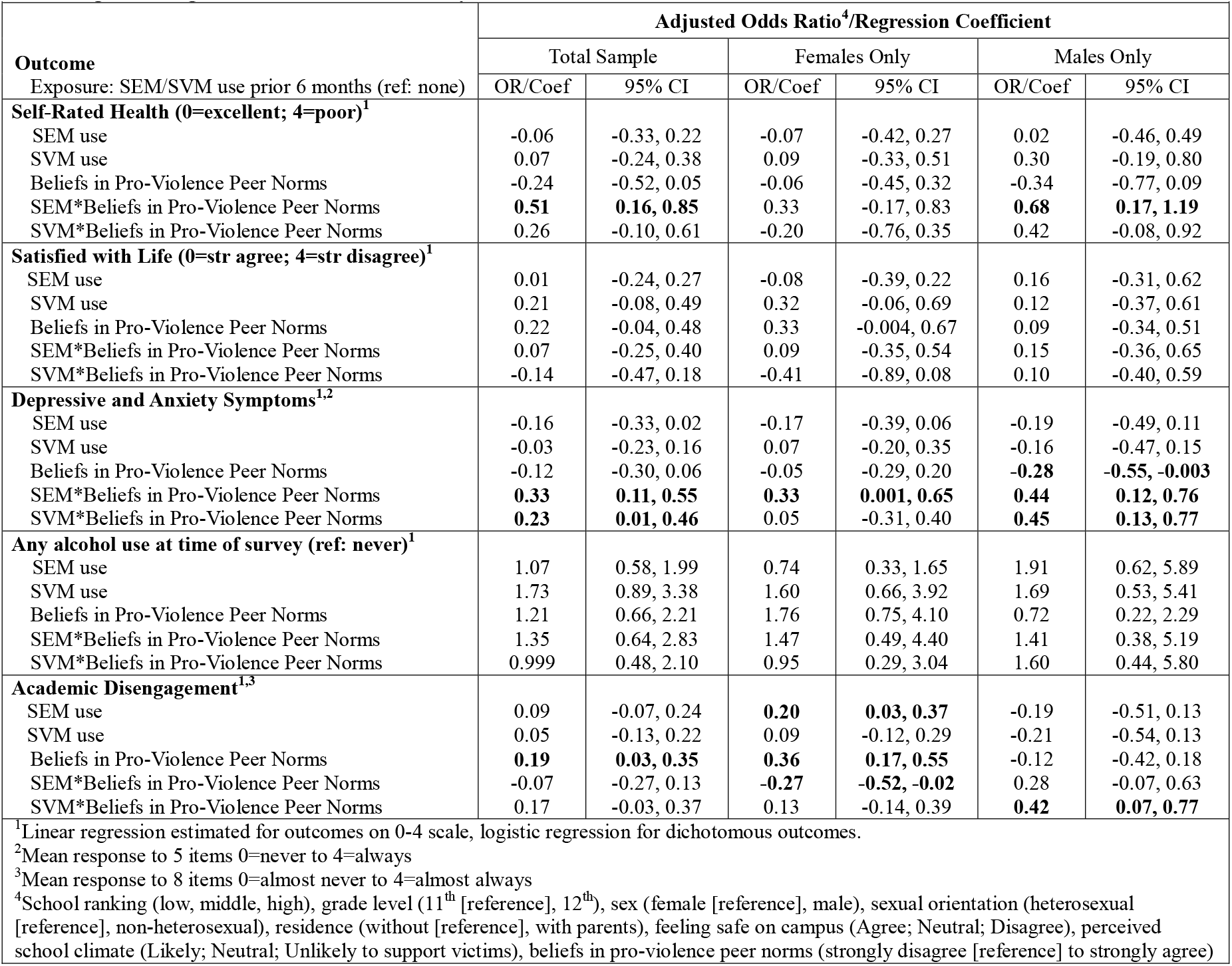
Adjusted Associations of Media Use with Health Outcomes, Including SEM-by-Belief and SVM-by-Belief Interactions, Students Attending Three High Schools in Ho Chi Minh City, Vietnam, 2023.

| Outcome<br>Exposure: SEM/SVM use prior 6 months (ref: none) | Adjusted Odds Ratio<sup>4</sup>/Regression Coefficient |  |  |  |  |  |
| --- | --- | --- | --- | --- | --- | --- |
|  | Total Sample |  | Females Only |  | Males Only |  |
|  | OR/Coef | 95% CI | OR/Coef | 95% CI | OR/Coef | 95% CI |
| **Self-Rated Health (0=excellent; 4=poor)<sup>1</sup>** |  |  |  |  |  |  |
| SEM use | -0.06 | -0.33, 0.22 | -0.07 | -0.42, 0.27 | 0.02 | -0.46, 0.49 |
| SVM use | 0.07 | -0.24, 0.38 | 0.09 | -0.33, 0.51 | 0.30 | -0.19, 0.80 |
| Beliefs in Pro-Violence Peer Norms | -0.24 | -0.52, 0.05 | -0.06 | -0.45, 0.32 | -0.34 | -0.77, 0.09 |
| SEM*Beliefs in Pro-Violence Peer Norms | **0.51** | **0.16, 0.85** | 0.33 | -0.17, 0.83 | **0.68** | **0.17, 1.19** |
| SVM*Beliefs in Pro-Violence Peer Norms | 0.26 | -0.10, 0.61 | -0.20 | -0.76, 0.35 | 0.42 | -0.08, 0.92 |
| **Satisfied with Life (0=str agree; 4=str disagree)<sup>1</sup>** |  |  |  |  |  |  |
| SEM use | 0.01 | -0.24, 0.27 | -0.08 | -0.39, 0.22 | 0.16 | -0.31, 0.62 |
| SVM use | 0.21 | -0.08, 0.49 | 0.32 | -0.06, 0.69 | 0.12 | -0.37, 0.61 |
| Beliefs in Pro-Violence Peer Norms | 0.22 | -0.04, 0.48 | 0.33 | -0.004, 0.67 | 0.09 | -0.34, 0.51 |
| SEM*Beliefs in Pro-Violence Peer Norms | 0.07 | -0.25, 0.40 | 0.09 | -0.35, 0.54 | 0.15 | -0.36, 0.65 |
| SVM*Beliefs in Pro-Violence Peer Norms | -0.14 | -0.47, 0.18 | -0.41 | -0.89, 0.08 | 0.10 | -0.40, 0.59 |
| **Depressive and Anxiety Symptoms<sup>1,2</sup>** |  |  |  |  |  |  |
| SEM use | -0.16 | -0.33, 0.02 | -0.17 | -0.39, 0.06 | -0.19 | -0.49, 0.11 |
| SVM use | -0.03 | -0.23, 0.16 | 0.07 | -0.20, 0.35 | -0.16 | -0.47, 0.15 |
| Beliefs in Pro-Violence Peer Norms | -0.12 | -0.30, 0.06 | -0.05 | -0.29, 0.20 | **-0.28** | **-0.55, -0.003** |
| SEM*Beliefs in Pro-Violence Peer Norms | **0.33** | **0.11, 0.55** | **0.33** | **0.001, 0.65** | **0.44** | **0.12, 0.76** |
| SVM*Beliefs in Pro-Violence Peer Norms | **0.23** | **0.01, 0.46** | 0.05 | -0.31, 0.40 | **0.45** | **0.13, 0.77** |
| **Any alcohol use at time of survey (ref: never)<sup>1</sup>** |  |  |  |  |  |  |
| SEM use | 1.07 | 0.58, 1.99 | 0.74 | 0.33, 1.65 | 1.91 | 0.62, 5.89 |
| SVM use | 1.73 | 0.89, 3.38 | 1.60 | 0.66, 3.92 | 1.69 | 0.53, 5.41 |
| Beliefs in Pro-Violence Peer Norms | 1.21 | 0.66, 2.21 | 1.76 | 0.75, 4.10 | 0.72 | 0.22, 2.29 |
| SEM*Beliefs in Pro-Violence Peer Norms | 1.35 | 0.64, 2.83 | 1.47 | 0.49, 4.40 | 1.41 | 0.38, 5.19 |
| SVM*Beliefs in Pro-Violence Peer Norms | 0.999 | 0.48, 2.10 | 0.95 | 0.29, 3.04 | 1.60 | 0.44, 5.80 |
| **Academic Disengagement<sup>1,3</sup>** |  |  |  |  |  |  |
| SEM use | 0.09 | -0.07, 0.24 | **0.20** | **0.03, 0.37** | -0.19 | -0.51, 0.13 |
| SVM use | 0.05 | -0.13, 0.22 | 0.09 | -0.12, 0.29 | -0.21 | -0.54, 0.13 |
| Beliefs in Pro-Violence Peer Norms | **0.19** | **0.03, 0.35** | **0.36** | **0.17, 0.55** | -0.12 | -0.42, 0.18 |
| SEM*Beliefs in Pro-Violence Peer Norms | -0.07 | -0.27, 0.13 | **-0.27** | **-0.52, -0.02** | 0.28 | -0.07, 0.63 |
| SVM*Beliefs in Pro-Violence Peer Norms | 0.17 | -0.03, 0.37 | 0.13 | -0.14, 0.39 | **0.42** | **0.07, 0.77** |

<sup>1</sup>Linear regression estimated for outcomes on 0-4 scale, logistic regression for dichotomous outcomes.
<sup>2</sup>Mean response to 5 items 0=never to 4=always
<sup>3</sup>Mean response to 8 items 0=almost never to 4=almost always
<sup>4</sup>School ranking (low, middle, high), grade level (11<sup>th</sup> [reference], 12<sup>th</sup>), sex (female [reference], male), sexual orientation (heterosexual [reference], non-heterosexual), residence (without [reference], with parents), feeling safe on campus (Agree; Neutral; Disagree), perceived school climate (Likely; Neutral; Unlikely to support victims), beliefs in pro-violence peer norms (strongly disagree [reference] to strongly agree)

## DISCUSSION

The use of sexually explicit media and sexually violent media among young people is a potential risk factor for many adverse social and health outcomes, including sexual misconduct perpetration and victimization.^4-6^ This risk may be due in part to increases in young people’s time spent using media and shifts to new media forms, which have increased access, especially among boys, to sexualized content that subordinates women and depicts violence against them and other vulnerable groups.^4^ Several shortcomings of this literature include unclear definitions of sexualized media; its assessment with few, unvalidated items that do not measure the modality (e.g., online) or category (e.g., violent content) of sexually explicit materials; potential bias of self-reported assessments; the multifactorial nature of SEM use and violence; cross-sectional designs that preclude causal inferences; a preponderance of studies with small, unrepresentative samples of white men from the U.S. that preclude cross-cultural generalizations; and atheoretical research on SEM use and violence, hindering comparisons of findings across studies.^51^

The present study addressed many of these limitations. It is the first study conducted in a large and diverse sample of male and female high-school students in Vietnam to collect detailed data on all major forms of sexual misconduct perpetration and victimization, other potential health outcomes, and refined measures of SEM and SVM use based on clear definitions and previously validated scales that showed high internal consistency in the current sample. The analysis was grounded in two theories for the effects of SEM and SVM use on sexual misconduct and well-being in high-school students: gendered aggression^5^ and moral incongruence.^25^ The analysis tested these theories by comparing SEM and SVM use levels among male and female students and associations of use with sexual misconduct perpetration and victimization (combining sexual harassment, sexual violence, physical dating violence, and stalking); perceived pro-violence peer norms; self-reported health, mental health, alcohol use, and school disengagement.

### Summary and Interpretation

Findings revealed that most (79.5% of) eleventh- and twelfth-grade students reported using any sexualized media in the prior six months. Nearly half (47.3%) reported using SEM only, and nearly one-third (32.2%) reported using any SVM. These use levels corroborate those among first-year undergraduate men in Hanoi, Vietnam, in which 86.2% reported any sexualized media use, 45.2% reported SEM use only, and 41.0% reported any SVM use in the same period.^7^ A study of exposure to sexually explicit internet material among students attending public high schools in four provinces in Vietnam showed that 84.1% of participants had been exposed, with near-universal use among male students (89.8%).^52^

Findings also confirmed expected differences in SEM and SVM use between male and female students. In the prior six months, female students more often reported non-use of any sexualized media (24.0% versus 15.4%), and male students more often reported SVM use (39.2% versus 27.3%). That said, SEM use was reported in just under half of female (48.7%) and male (45.5%) students. Prior studies in Vietnam have not differentiated the type of sexualized media used by high-school students,^52^ so the present study exposed questions about whether boys’ greater SVM use had implications for gendered aggression models of sexual misconduct perpetration and victimization.

Findings supported a general aggression model in the pooled sample^16^ (hypothesis 1) and partially supported a gendered aggression model in stratified samples (hypothesis 2). In the pooled sample, SVM use was positively associated with any sexual misconduct and any sexual violence perpetration and victimization, and SEM use was positively associated with sexual misconduct victimization. Supporting a gendered aggression model (hypothesis 2), positive relationships of SEM use with sexual misconduct victimization and SVM use with sexual violence victimization were observed only among girls. Thus, female users may learn, activate, and apply in their daily lives scripts in sexualized media that depict the objectification and physical violence of women. Prevailing gender norms among young people in Vietnam that normalize women’s compliance and men’s dominance, violence, and entitlement to sex may reinforce observed gendered sexual scripts in media.^31,34^

That said, a positive relationship between SVM use and sexual misconduct victimization was also observed among boys. In Vietnam, the gendered scripts learned by some male SVM users may align with experiences of emotional, physical, and sexual violence in childhood,^37^ including corporal punishment by male adult relatives^53^ and unwanted sexual attention and coercion by school staff.^8^ In parts of Vietnam, ethnographic research has suggested that corporal punishment by senior male relatives pushes the “…beaten boy into an inferior (feminine) position” (p. 336).^53^ Thus, the violent scripts learned in SVM may reinforce scripts learned from experiences of violence in childhood, making male high-school student SVM users more vulnerable to sexual misconduct victimization in their daily lives.

Finally, SVM use was positively associated with sexual misconduct perpetration among male and female students. This finding for boys corroborates a gendered aggression model (hypothesis 2) and findings from a prior study of first-year undergraduate men in Vietnam; in that study, any SVM use in the prior six months was associated with higher adjusted odds of any non-contact sexually violent behavior and contact sexually violent behavior using physical and non-physical tactics.^7^ Both findings support the possibility that male SVM users in Vietnam observe gendered sexual scripts in media amidst prevailing gender norms that normalize masculine dominance and violence toward others. However, the finding that SVM use was associated with misconduct perpetration among girls was unexpected. Here, a general aggression model may apply to some girls who use SVM in Vietnam. Alternatively, the motivation for perpetration among girls may be self-defense. According to young people in Vietnam, girls should show “fierce resistance” when sexual violence is initiated against them to confirm non-consent.^31,34,54^ Finally, the type and target of misconduct perpetration may differ for girls and boys; however, the sample sizes of female and male students were not large enough to analyze each type of sexual misconduct perpetration separately, and data on the target of violence was not collected.

Findings supported a moral incongruence model for the associations of SEM use and SVM use especially with boys’ health and well-being (hypotheses 3-5). Among boys, compared to non-use, SEM and SVM use were associated with poorer self-rated health and higher adjusted odds of alcohol use. These relationships may be bidirectional,^25^ and causality cannot be inferred from the present cross-sectional study. Still, questions about SEM and SVM use were asked regarding “the prior six months,” and questions about self-rated health and alcohol use were asked regarding the time of the survey; thus, findings support the idea that prior use of sexualized media may be linked with subsequent well-being outcomes.^55^ Models with interaction terms revealed that boys who used SEM had worse self-rated health and worse mental health when they believed more strongly that peers supported violence against women. Similarly, boys who used SVM had worse mental health and higher academic disengagement when they believed more strongly that peers supported violence against women. These findings suggest that boys who use SEM and SVM and who more strongly perceive pro-violence norms among peers may feel the strongest cognitive dissonance between their own moral values and use practice. Among high-school girls, largely as expected, beliefs about pro-violence peer norms did not alter associations of SVM use with well-being outcomes and either did not alter or inconsistently altered the association of SEM use with well-being outcomes.

### Limitations and Strengths

Some non-participation may have occurred due to data collection in May, the examination period for high schools in Ho Chi Minh city. Still, the participation rate (75.0%) far exceeded rates in surveys of undergraduates in the U.S. (20.4%).^56^ Also, higher levels of missingness in later modules about sexual violence victimization (16%-23%) than in earlier modules about sexual harassment (11%-14%) may have been due to respondent burden, the greater sensitivity of questions about sexual violence, and some duplicate records for students who lost internet connectivity and re-entered REDCap with a new ID to retake the survey. Still, levels of missingness in our study and surveys of undergraduates in the U.S. were similar,^57^ and estimates of sexual misconduct are similar across different response rates.^58^ Finally, despite this study’s relatively large sample size,^24^ it still was too small to analyze forms of sexual misconduct separately and “frequent” versus “any” SVM use separately, which is relevant for aggression theory. Still, research with undergraduate men in Vietnam showed that adjusted odds of sexually violent behavior were similar for men reporting any and at least monthly SVM use.^7^

The study design addressed several limitations of prior studies,^51^ lending credibility to the findings. The survey collected detailed data on multiple forms of sexual misconduct victimization and perpetration. Measures relied on previously validated scales^1,39^ that had good internal consistency in the present sample.^8^ The survey included contextualized demographic questions from prior surveys of young people in Vietnam^35^ and a module on SEM/SVM use that was self-administered online among young men in Vietnam.^7^ The anonymous online self-administered survey format likely increased participation rates compared to similar surveys of students in the U.S.^56^ and disclosures to sensitive questions.

### Implications

Findings have important implications for future research and policy. First, a national high-school climate survey with a module on media use is warranted. Second, survey experiments are recommended to understand how design elements affect participation and completion. Randomizing the number and order of survey modules; sampling strategy; delivery mode; compensation level and type; and/or number, frequency, and format of participation and completion reminders would help to identify the design elements that optimize data quality in future surveys.

Third, longitudinal studies could confirm or disconfirm the cross-sectional findings among high-school students presented here. Fourth, a multi-arm randomized controlled trial would be useful to test the effects of phone-free school environments, alone and with evidence-based educational programming with teachers, parents, and students, on the health and behavioral harms of sexualized media use among students. Finally, findings should be disseminated locally to raise awareness among policymakers, school leaders, and parents about the potential benefits of contextualized school policies and programs to reduce harmful media use as one mechanism to improve student well-being.

## CONCLUSION

In this first comprehensive climate survey of high-school students in Vietnam, sexually explicit (47%) and sexually violent (32%) media use was common and had gender-differentiated associations with poorer self-rated health, alcohol use, and violence victimization and perpetration. Future programming may involve phone-free school days and evidence-based education to staff, parents, and students to reduce the specific harms of sexualized media on young people.

## Data Availability

All data produced in the present study are available upon reasonable request to the corresponding author.

https://www.medrxiv.org/content/10.1101/2025.01.28.25321240v1

## CRediT authorship statement

<u>Kathryn M. Yount</u>: Conceptualization, Funding acquisition, Methodology, Project administration, Supervision, Visualization, Writing – original draft, Writing – review and editing

<u>Meghan Macaulay</u>: Formal analysis, Methodology, Verification, Writing – review & editing

<u>Kim Tu Tran</u>: Data curation, Funding acquisition, Investigation, Methodology, Project administration, Resources, Supervision, Writing – review & editing

## Acknowledgments

The authors thank the participants, without whom this project would not have been possible.

## Data Statement

This study was conducted in a small area, which increases the risk of identification; therefore, we are not making the data publicly available. The corresponding author can provide the data upon reasonable request.

